# A novel monocyte-based biomarker of cardiovascular risk: comparison with traditional cardiovascular risk calculators

**DOI:** 10.1101/2024.07.12.24310323

**Authors:** Fatemah Almarri, Shuaihua Qiao, Jiale Gao, Li’na Kang, Soundrie Padayachee, Biao Xu, Ashish Patel, Albert Ferro

**Affiliations:** School of Cardiovascular and Metabolic Medicine and Sciences, British Heart Foundation Centre of Research Excellence, King’s College London, London, UK; Department of Cardiology, Nanjing Drum Tower Hospital, The Affiliated Hospital of Nanjing, University Medical School, Nanjing University, Nanjing, China; Department of Ultrasonic Angiology, Guy’s & St Thomas’ Hospitals, London, UK

**Author notes:** Contributed equally to this work (Fatemah Almarri, Shuaihua Qiao). Co-senior authors (Ashish Patel, Albert Ferro). <u>**Correspondence:**</u> Professor Albert Ferro, King’s College London (Waterloo Campus), 3.07 Franklin-Wilkins Building, 150 Stamford Street, London SE1 9NH, UK.

**Keywords:** monocytes, monocyte-platelet aggregates, atherosclerosis, cardiovascular risk, biomarkers

## Abstract

**Background:** Traditional cardiovascular risk calculators estimate population-level risk but perform less reliably at the individual level and do not capture biological processes that underlie early atherogenesis. Monocyte activation and platelet–monocyte interactions contribute to early atherogenesis, yet their potential as biomarkers of silent disease in asymptomatic adults has not been defined. The aim of this study was to investigate their utility in prediction of early atherosclerosis in asymptomatic subjects, in comparison to traditional cardiovascular risk calculators.

**Methods:** Asymptomatic adults in a discovery cohort (n=39) underwent flow cytometric profiling of monocyte subsets and monocyte–platelet aggregates (MPA), together with carotid ultrasonography to assess carotid intima–media thickness (cIMT) and plaque. From these data, we derived a composite biomarker, the Monocyte Atherosclerotic Risk Score (MARS). Cardiovascular risk was calculated using QRISK3. An independent validation cohort of clinically healthy subjects (n=151) attending the Physical Examination Centre at Drum Tower Hospital, Nanjing, China, underwent identical biomarker and imaging assessments, with cardiovascular risk calculated using China-PAR.

**Results:** In the discovery cohort, MARS showed a strong association with cIMT (r²=0.87, P<0.0001), substantially outperforming QRISK3 (r²=0.30, P=0.003). MARS predicted high-risk cIMT (AUC 0.93, P=0.0001) and carotid plaque (AUC 0.94, P=0.0022), whereas QRISK3 did not. In the validation cohort, MARS again discriminated individuals with carotid plaque (AUC 0.81, P<0.0001), while China-PAR showed no significant predictive ability.

**Conclusions:** The MARS biomarker generated from flow cytometric profiling detects silent atherosclerosis with higher accuracy than QRISK3 or China-PAR in two independent and ethnically different asymptomatic populations. These findings support further evaluation of MARS as a scalable blood-based tool for identifying individuals who may benefit from targeted imaging and preventive strategies.

## INTRODUCTION

Atherosclerotic cardiovascular disease and its complications (including myocardial infarction, stroke and peripheral arterial disease) remains the leading global cause of death and disability despite significant advances in prevention and treatment. ^1^ A major challenge is the identification of individuals with subclinical asymptomatic disease. A recent coronary CT angiography study of more than 25 000 adults without known coronary disease showed that 42.1% had detectable atherosclerosis, including significant (≥50%) stenosis in 5.2% and left main, proximal left anterior descending (LAD) or three-vessel disease in 1.9% of subjects, underscoring the high burden of silent disease. ^2^ However, routine imaging of asymptomatic populations is not feasible, highlighting the need for scalable biomarkers capable of identifying individuals with early, clinically silent pathology.

Current cardiovascular risk calculators include QRISK3, PREVENT and the European Society of Cardiology cardiovascular risk calculator, all of which estimate the likelihood of developing cardiovascular disease over a fixed time period or over the subject’s lifetime based on their risk factor profile.^3–5^ Although these calculators perform well at a population level, they do not necessarily give an accurate prediction of risk in a given individual. For example, data from UK Biobank involving 233,233 women and 170,137 men showed that QRISK3 only moderately discriminated for participants (Harrell’s C-index 0.722 in women and 0.697 in men), with performance declining by age; in older patients QRISK3 systematically overpredicted cardiovascular risk by as much as 20%.^6^ Similarly, a study by Li et al using QRISK3 as an exemplar demonstrated that risk prediction models based on routinely collected health data perform adequately for populations but with substantially greater uncertainty for individuals.^7^ These limitations highlight the need for improved approaches to estimating cardiovascular risk at an individual level, including novel biomarkers that more directly reflect underlying disease biology.

Recent years have seen increasing interest in the role of inflammation in the pathophysiology of atherosclerosis, with monocytes recognised as key cellular contributors to early plaque development.^8^ Circulating CD16-expressing monocytes subsets and monocyte-platelet aggregates (MPAs) are elevated in patients with symptomatic atherosclerotic disease including coronary heart disease, stroke and peripheral arterial disease, and are associated with disease progression and cardiovascular events.^9–14^ Despite this, their utility as biomarkers of early, asymptomatic atherosclerosis has not previously been evaluated.

In the present study, we examined whether circulating monocyte phenotype and MPA levels could identify the presence and extent of subclinical atherosclerosis. We hypothesised that these biomarkers would prove superior to standard cardiovascular risk calculators in such prediction.

## METHODS

### Subjects

#### Discovery Cohort

The study was approved by HRA and Health and Care Research Wales (research ethics committee reference number 21/NE/0189). All subjects gave written informed consent. For the initial discovery analysis, asymptomatic adults (n=39) were recruited either by advertisement or sequentially from patients attending the Hypertension Clinics at Guy’s and St Thomas’ Hospitals, London, UK. Exclusion criteria were: age<18 years, history of cardiovascular disease (other than hypertension), significant co-morbidities, ingestion of any medication within the preceding 2 weeks, pregnancy, or inability/unwillingness to provide informed consent. All participants underwent a clinical history, physical examination and electrocardiography to exclude overt cardiovascular or other systemic disease. Blood pressure was measured in the sitting position, after a resting period of 5 min, using a validated oscillometric device (Omron IntelliIT, Omron Corporation, Japan), according to the recommendations of the British and Irish Hypertension Society.^15^ Three readings were obtained at two-minute intervals, and the mean of the second and third values was used for analysis. Venous blood (50 mL) was then collected for full blood count and biochemistry (renal, liver, bone, thyroid and lipid profiles, plasma renin, aldosterone and metanephrines), these being undertaken by Synnovis Laboratories (St Thomas’ Hospital, London, UK), and additionally flow cytometric analysis was performed as described below. Within one month of venesection, participants underwent carotid ultrasonography to assess carotid intima–media thickness (cIMT) and the presence or absence of plaque disease. Ten-year cardiovascular risk was estimated using QRISK3 (https://qrisk.org).

#### Validation Cohort

For the subsequent validation study, the study was approved by the Medical Ethics Committee of Nanjing Drum Tower Hospital, Medical School of Nanjing University, Nanjing, China (ethics committee reference 2025-0115-02). Clinically healthy adults (n=151) were recruited from the Physical Examination Centre at Drum Tower Hospital, Nanjing, where routine medical evaluations are performed for employment or health screening. Exclusion criteria and screening procedures were identical to those used in the discovery cohort. Blood sampling and carotid ultrasonography were performed using the same protocols, with the exception that haematological and biochemical analyses were conducted in the hospital’s pathology laboratories. Ten-year cardiovascular risk was calculated using the China-PAR model (https://doi.org/10.3760/cma.j.issn.0253-9624.2019.01.004), the clinical standard for China.

### Flow cytometry of peripheral blood mononuclear cells

Peripheral blood was processed within 15-30 minutes of venesection. Whole blood (100µl) was incubated with 20µl Human BD Fc-block (BD Pharmingen^TM^) for 10 min on ice to minimise non-specific antibody binding. Samples where then stained in the dark at 4°C for 30min with 5µl of the following fluorochrome-conjugated antibodies: PE-labelled anti- CD14, FITC-labelled anti- CD16, APC-labelled anti-CD42b, BV421-labelled anti-CD200R, BV711-labelled anti-CD163, Alexa Fluor 647-labelled anti-CCR2 (CD192), and PE- Cy7 anti- HLA-DR (all BD Pharmingen). Following staining, red blood cells were lysed using Pharm Lyse (BD Biosciences) and washed using flow cytometry buffer (PBS, 0.5% [w/v] BSA, 2mM EDTA). Stained cells were fixed in 4% paraformaldehyde and analysed within 72 h on a MACSQuant Flow Cytometer (Miltenyi Biotec). Fluorescence-minus-one (FMO) controls were used to define positive gates for each marker. . Data analysis was performed using FlowJo v10 (FlowJo LLC) by a researcher blinded to clinical information. Peripheral blood mononuclear cells were identified by forward and side scatter and granulocytes and doublets excluded. Classical (CD14^++^CD16^−^), intermediate (CD14^++^CD16^+^), and non-classical (CD14^low^CD16^+^) monocyte subsets were gated based on CD14 and CD16 expression. Monocyte–platelet aggregates (MPA) were defined as CD14^+^CD42b^+^ events. Extracellular epitope expression was quantified as the percentage of cells positive for each marker within the monocyte population.

### Carotid Duplex Ultrasonography

cIMT was assessed using standardised protocols. All scans were performed by accredited vascular scientists blinded to biomarker data. The common carotid artery was imaged in longitudinal section with the ultrasound beam perpendicular to the posterior wall to optimise delineation of the intima–media complex. Electrocardiography-gated images were obtained at end-diastole (R-wave). The common carotid artery was imaged in anterior, posterior and lateral projections and measurements were taken at least 10 mm proximal to the carotid bifurcation. Semi-automated edge detection software (Qlab; Philips Healthcare) was used to derive cIMT values from the posterior wall. For each side, the maximum cIMT was defined as the highest value from the average of 200 measurements across a 1 cm arterial segment. Right and left cIMT values were recorded separately. The presence of carotid plaque was defined as a focal lesion with thickness >1.5 mm, measured from the media–adventitia interface to the intima–lumen boundary.

cIMT was interpreted relative to age- and sex-matched adjusted reference standards for cardiovascular disease risk assessment.^16^

### Statistics

Data were analysed using GraphPad Prism version 10 (GraphPad Software, San Diego, California, USA). Continuous variables are presented as mean ± standard deviation for normally distributed data, or median with interquartile range for non-normally distributed data. Normality was assessed using Shapiro–Wilk tests where appropriate. Associations between continuous variables were evaluated using linear regression. The predictive performance of monocyte-derived indices and cardiovascular risk calculators for high-risk cIMT and for the presence of carotid plaque was assessed using receiver operating characteristic (ROC) curve analysis. Discrimination was summarised using the area under the curve (C-index), with 95% confidence intervals. Comparisons of C-index values were descriptive; inferential comparisons were not performed given the sample size. For all analyses, statistical significance was taken as P<0.05.

## RESULTS

### 1. DISCOVERY STUDY

#### Subject characteristics

Baseline characteristics of participants recruited for the discovery study are shown in Tables 1 and 2. Participants recruited from the hypertension clinic had higher blood pressure and were more likely to be receiving cardiovascular medications than those recruited by advertisement. Other demographic variables, including age, sex, smoking status, haematological indices and biochemical parameters, did not differ significantly between groups. For subsequent analyses, participants from both recruitment sources were pooled and analysed as a single cohort

**Table 1.** Demographics of subjects recruited from the hypertension clinic.

|  |  |
| --- | --- |
| **Number (male / female)** | 14/12 |
| **Age (years)** | 41.1±16.3 |
| **Smokers** | 14 |
| **Systolic blood pressure (mmHg)** | 141.6±10.5 |
| **Diastolic blood pressure (mmHg)** | 88.7±10.3 |
| **Creatinine (μmol/L)** | 76.4±15.9 |
| **HbA<sub>1c</sub> (%)** | 5.77±0.10 |
| **Total cholesterol (mmol/L)** | 4.95±1.06 |
| **LDL-cholesterol (mmol/L)** | 3.04±0.93 |
| **HDL-cholesterol (mmol/L)** | 1.39±0.42 |
| **Triglycerides (mmol/L)** | 1.42±0.96 |
| **Haemoglobin (g/L)** | 140.0±12.8 |
| **Total white cell count (× 10<sup>9</sup>/L)** | 6.14±1.41 |
| **Neutrophil count (× 10<sup>9</sup>/L)** | 3.73±1.20 |
| **Monocyte count (× 10<sup>9</sup>/L)** | 0.48±0.15 |
| **Lymphocyte count (× 10<sup>9</sup>/L)** | 1.67±0.47 |
| **Platelet count (× 10<sup>9</sup>/L)** | 254.0±13.8 |
| **Medications** |  |
| - Beta blockers | 3 |
| - Alpha blockers | 2 |
| - Calcium channel blockers | 8 |
| - Renin-angiotensin system inhibitors | 14 |
| - Aspirin | 2 |
| - Diuretics | 6 |
| - Statins | 8 |

**Table 2.** Demographics of subjects recruited by advertisement.

|  |  |
| --- | --- |
| **Number (male / female)** | 5/8 |
| **Age (years)** | 39.6±11.7 |
| **Smokers** | 6 |
| **Systolic blood pressure (mmHg)** | 123.9±7.1 |
| **Diastolic blood pressure (mmHg)** | 78.8±2.1 |
| **Creatinine (μmol/L)** | 75.7±19.2 |
| **HbA<sub>1c</sub> (%)** | 5.63±0.25 |
| **Total cholesterol (mmol/L)** | 5.38±0.73 |
| **LDL-cholesterol (mmol/L)** | 3.47±0.72 |
| **HDL-cholesterol (mmol/L)** | 1.44±0.23 |
| **Triglycerides (mmol/L)** | 1.07±0.34 |
| **Haemoglobin (g/L)** | 129.4±16.0 |
| **Total white cell count (× 10<sup>9</sup>/L)** | 5.44±1.69 |
| **Neutrophil count (× 10<sup>9</sup>/L)** | 3.19±1.26 |
| **Monocyte count (× 10<sup>9</sup>/L)** | 0.45±0.14 |
| **Lymphocyte count (× 10<sup>9</sup>/L)** | 2.07±0.61 |
| **Platelet count (× 10<sup>9</sup>/L)** | 240.1±54.0 |

#### Carotid ultrasonography

Of the 39 participants in the discovery cohort, 10 were found to have either unilateral or bilateral carotid plaque disease on Duplex ultrasonography. Measurements of cIMT were highly concordant between the left and right common carotid arteries (Figure 2A), with minimal bias demonstrated on Bland–Altman analysis (Figure 2B). Accordingly, the mean of left and right cIMT values was used for all subsequent analyses..

**Figure 1.**
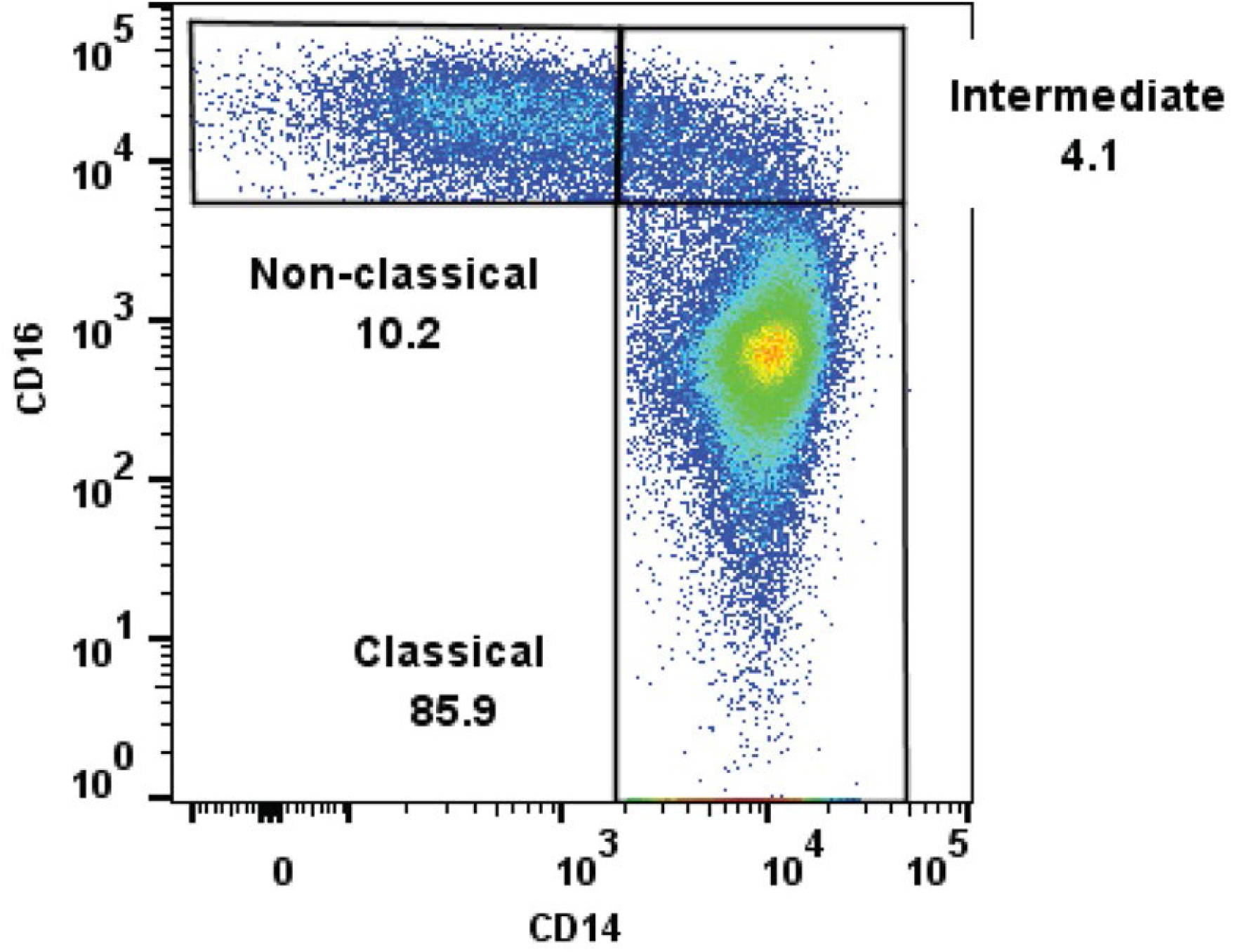
Gating for CM, NCM, and IM. Gating for monocyte subsets (classical, intermediate and non-classical) according to CD14 and CD16 expression and using respective fluorescence minus one (FMO) control samples.

**Figure 2.**
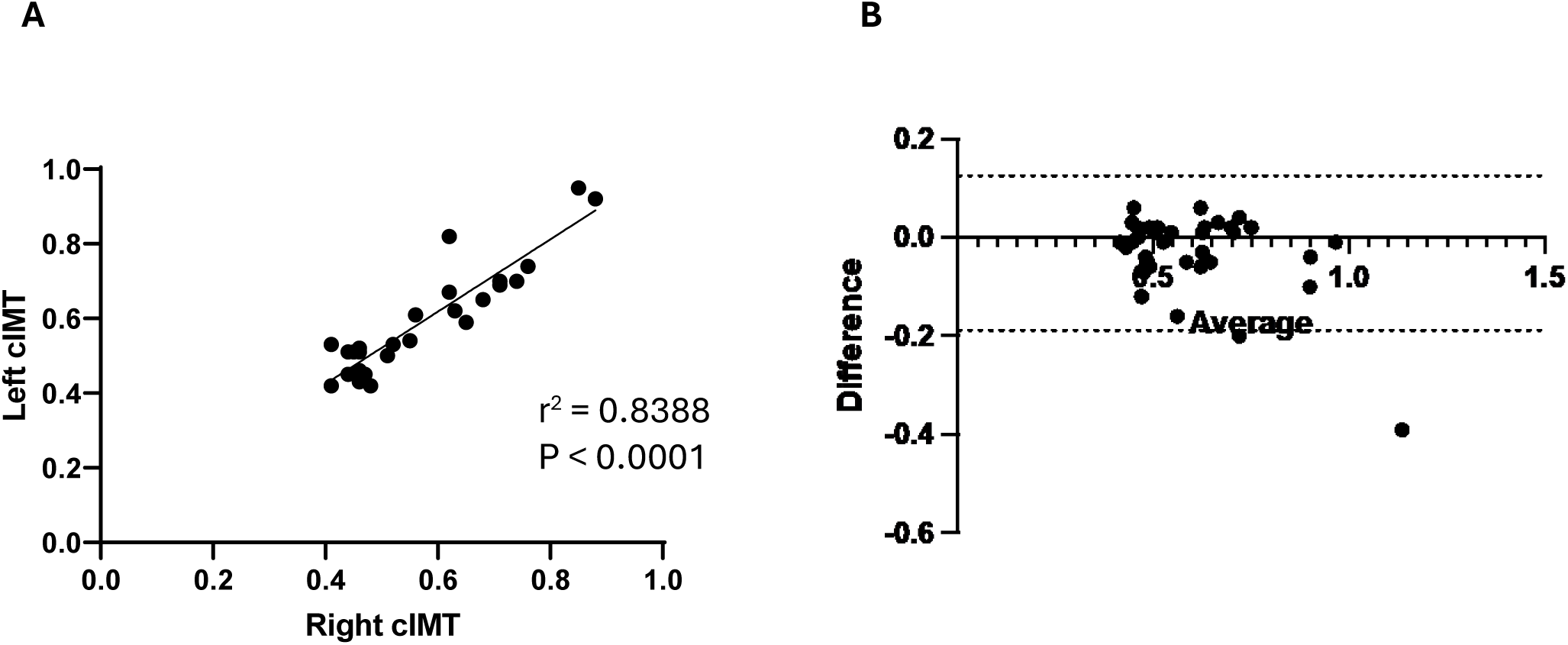
Agreement between left and right cIMT in the discovery cohort. **A,** Linear correlation analysis of left vs right cIMT. **B,** Bland-Altman plot of difference vs average of left and right cIMT; the dotted lines represent the 95% limits of agreement.

#### Relationship between monocyte phenotype, monocyte–platelet aggregates (MPAs) and cIMT

The proportion of classical (CD14^++^CD16^-^) monocytes was inversely associated with mean cIMT, whereas both intermediate (CD14^++^CD16^+^) and non-classical (CD14^low^CD16^+^) monocyte subsets showed strong positive associations with cIMT (Figure 3A–C). MPA levels also positively correlated with cIMT (Figure 3D).

**Figure 3.**
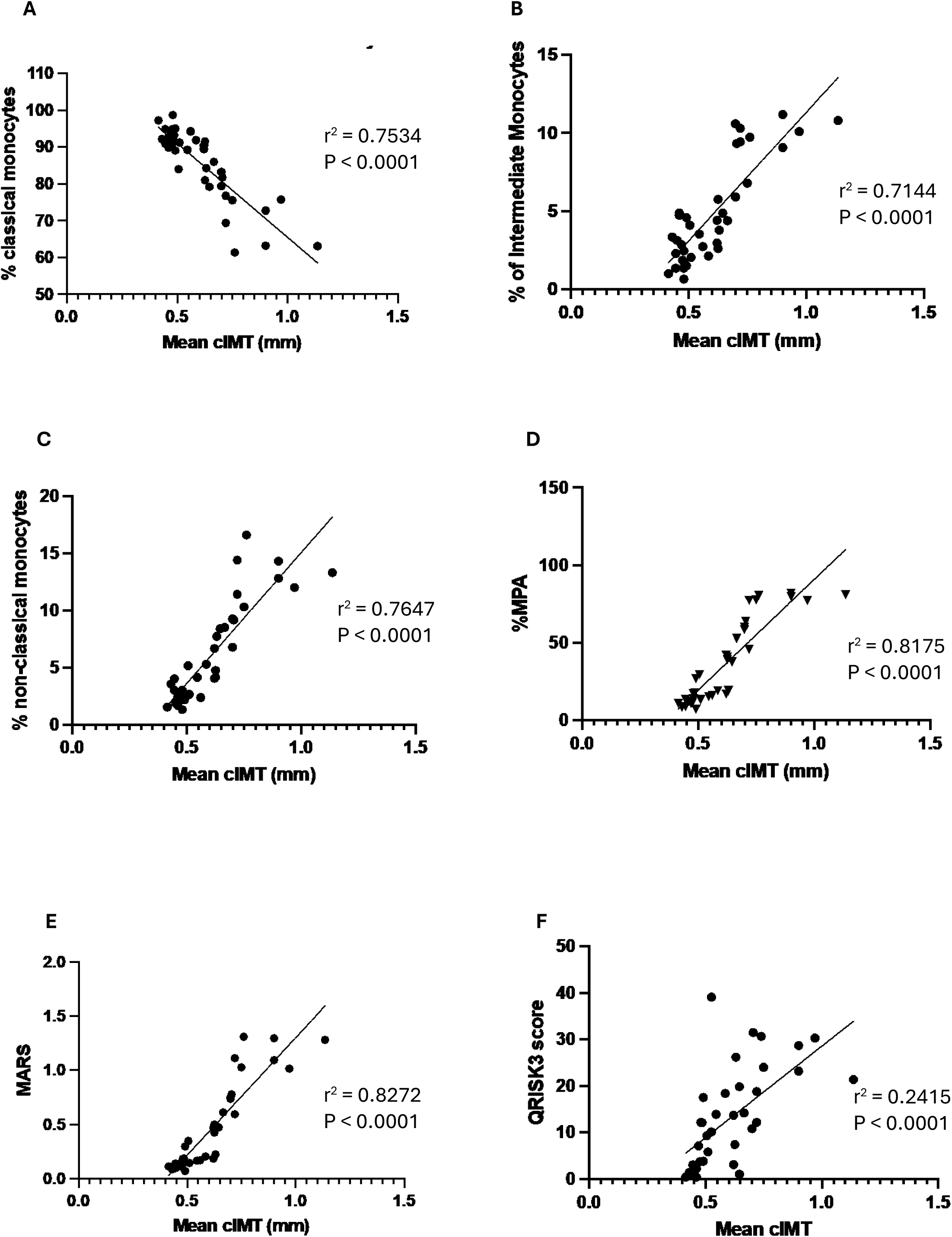
Linear regression analyses of the relationship between mean cIMT and monocyte indices or QRISK3 score in the discovery cohort. Relationships are shown between mean cIMT and classical (**A**), intermediate (**B**) and non-classical (**C**) monocytes, MPAs (**D**), MARS (**E**) and QRISK3 score (**F**).

To integrate these findings, we derived a novel composite index, the Monocyte Atherosclerotic Risk Score (MARS), defined as the ratio of MPA to classical monocytes. MARS demonstrated the strongest association with cIMT among all monocyte-related variables examined (Figure 3E)

QRISK3 scores were also positively associated with with cIMT (Figure 3F); however, the strength of this association was substantially weaker than that seen for monocyte subsets, MPA or MARS (as reflected by lower a r^2^ value and greater inter-individual variability).

#### Relationship between MARS and QRISK3

Although MARS and its component variables were significantly correlated with QRISK3, these relationships were modest, with low r² values indicating limited shared variance (Figure 4A–E). This suggests that MARS captures biological information that is not fully reflected by traditional risk factor-based models.

**Figure 4.**
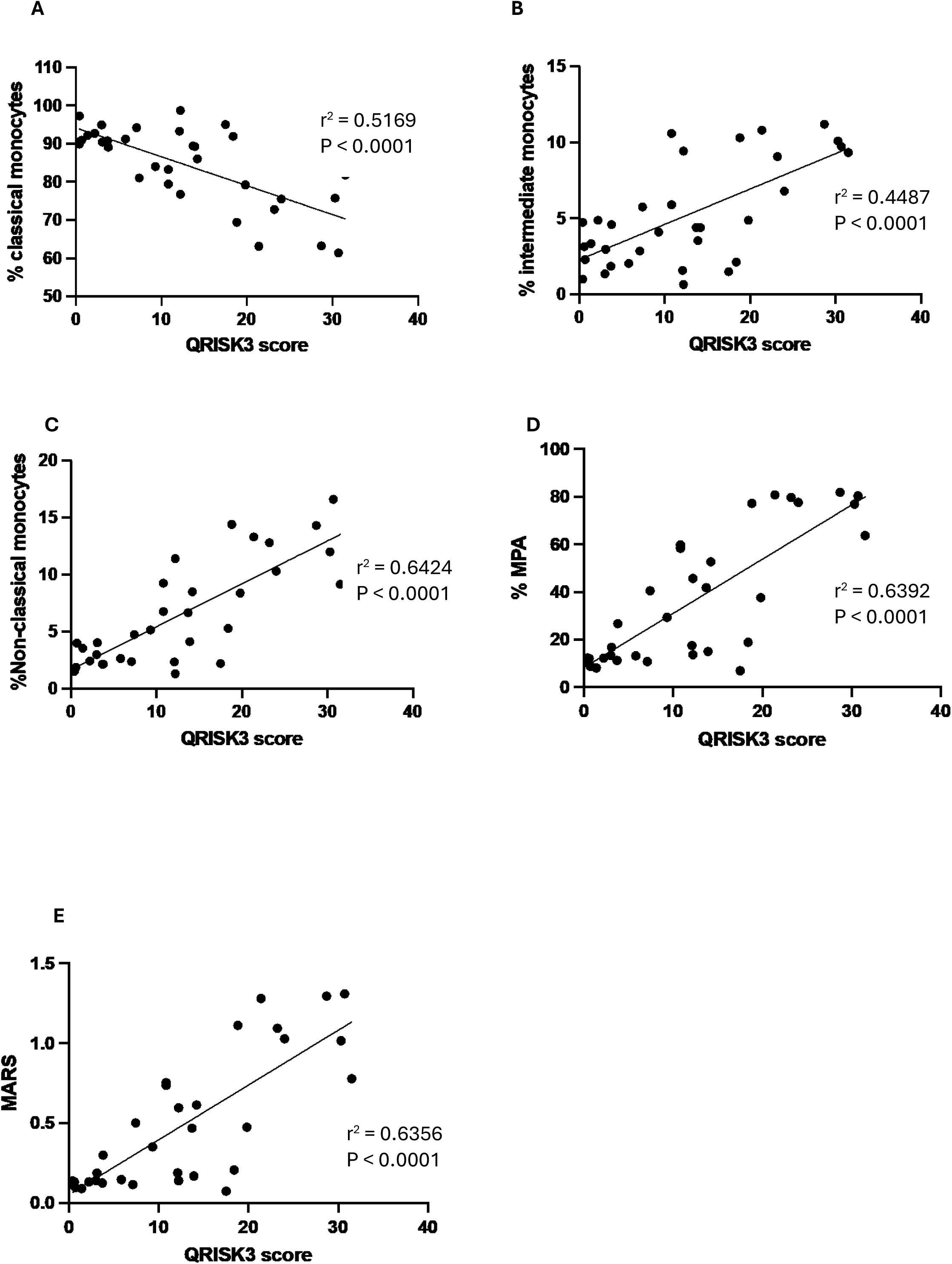
Linear regression analyses of relationship between QRISK3 score and monocyte indices in the discovery cohort. Relationships are shown between QRISK3 score and classical (**A**), intermediate (**B**) and non-classical (**C**) monocytes, MPAs (**D**) and MARS (**E**).

#### Associations of additional monocyte markers with cIMT

We also wanted to delineate the relationship between cIMT and monocytic expression of a number of other markers, (CD200R, CD163, CD192 and HLA-DR) which have been reported to be associated with atherosclerosis.^17–20^. Expression of CD200R on circulating monocytes was inversely associated with cIMT, whereas expression of CD163, CCR2 (CD192) and HLA-DR showed positive associations with cIMT (Figure 5A–D). These relationships were non-linear and weaker than those observed for MARS.

**Figure 5.**
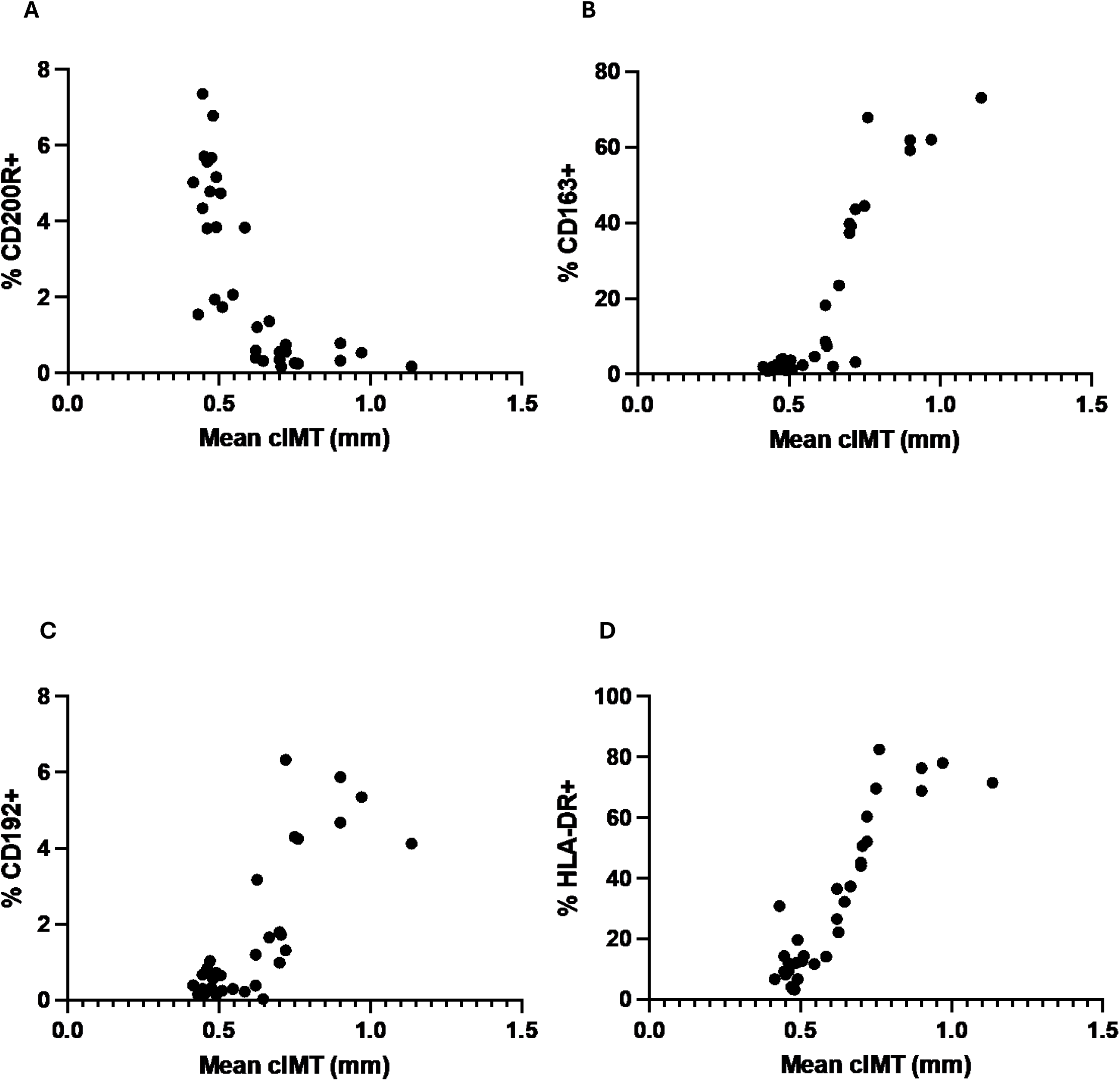
Relationship between mean cIMT and other monocyte indices in the discovery cohort. Relationships are shown between mean cIMT and monocytic expression of CD200R (**A**), CD163 (**B**), CD192 (**C**) and HLA-DR (**D**).

#### Prediction of high-risk cIMT

Using age- and sex-adjusted reference values, participants were classified as having high-risk cIMT if values were at or above the 75th percentile. ^21^ ROC analysis demonstrated that MARS robustly discriminated individuals with high-risk cIMT (C-index 0.93, P=0.0001; Figure 6A). QRISK3 showed weaker discrimination (Figure 6B). Combining MARS with QRISK3 did not improve predictive performance compared with MARS alone. Similarly, inclusion of the additional monocyte surface markers (either individually or in combination) did not enhance discriminatory ability beyond that achieved by MARS (Table 3).

**Figure 6.**
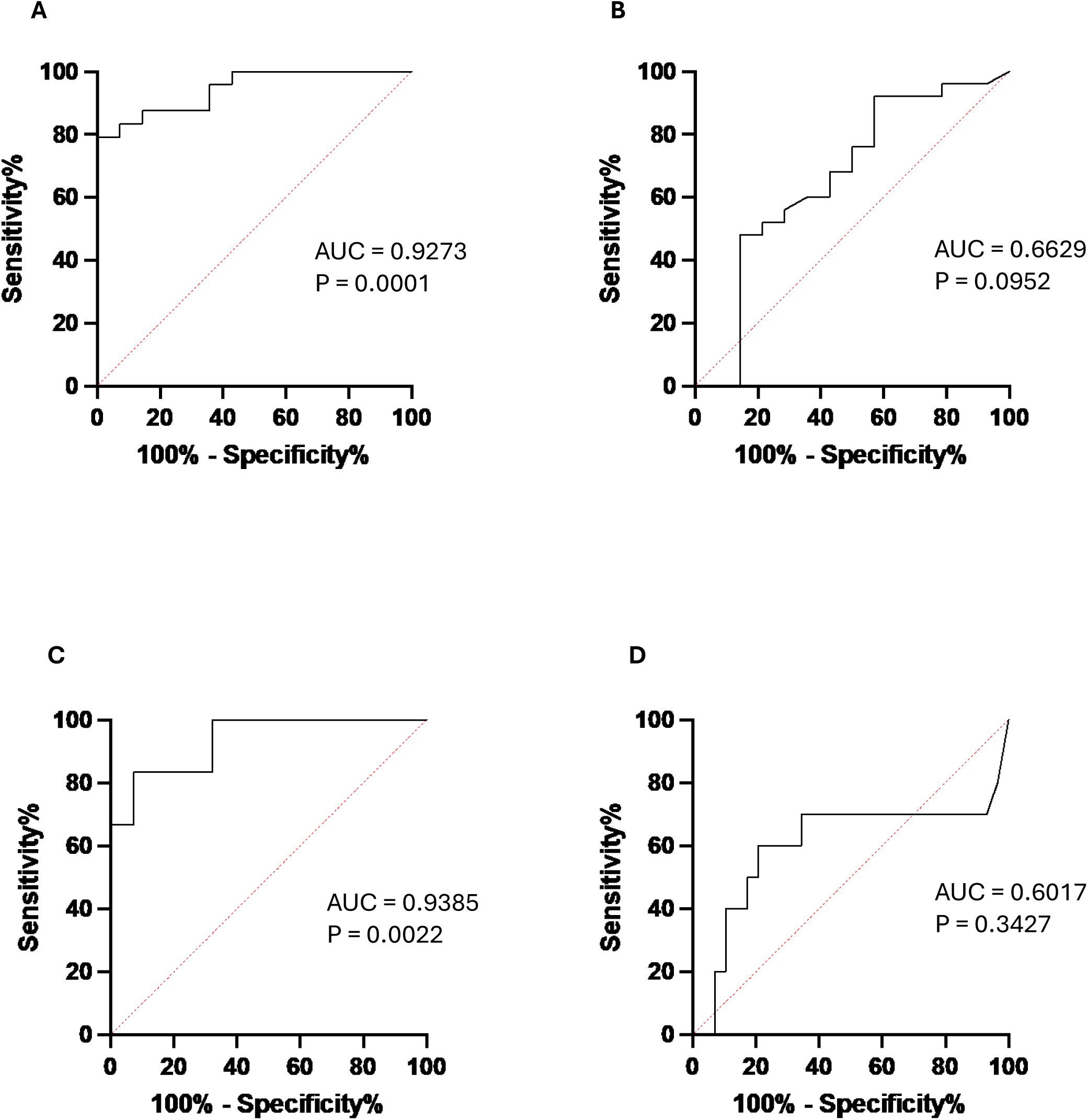
Receiver operating characteristic (ROC) curve analysis of predictive ability of MARS and QRISK3 score for carotid disease in the discovery cohort. ROC curves are shown for ability of MARS to predict high risk cIMT (**A**) or presence of plaque (**C**), and of QRISK3 score to predict high risk cIMT (**B**) or presence of plaque (**D**).

**Table 3.** C-index for prediction of high risk cIMT: MARS alone and in combination with QRISK3 score or with other monocyte biomarkers.

|  | **C-index (95% CI)** | **P value** |
| --- | --- | --- |
| **MARS** | 0.9273 (0.8414-1.000) | 0.0001 |
| **MARS and QRISK3** | 0.8409 (0.7053-0.9766) | 0.0023 |
| **MARS and CD200R** | 0.8909 (0.7815-1.000) | 0.0005 |
| **MARS and CD163** | 0.9136 (0.8151-1.000) | 0.0002 |
| **MARS and CD192** | 0.7955 (0.6417-0.9492) | 0.0082 |
| **MARS and HLA-DR** | 0.8909 (0.7748-1.000) | 0.0005 |
| **MARS and CD200R and CD163** | 0.9136 (0.8146-1.000) | 0.0002 |
| **MARS and CD200R and CD192** | 0.8500 (0.7185-0.9815) | 0.0017 |
| **MARS and CD200R and HLA-DR** | 0.8773 (0.7592-0.9954) | 0.0007 |
| **MARS and CD163 and CD192** | 0.8455 (0.7112-0.9798) | 0.0020 |
| **MARS and CD163 and HLA-DR** | 0.9273 (0.8392-1.000) | 0.0001 |
| **MARS and CD192 and HLA-DR** | 0.8545 (0.7243-0.9848) | 0.0015 |
| **MARS and CD200R and CD163 and CD192** | 0.8682 (0.7453-0.9910) | 0.0010 |
| **MARS and CD200R and CD163 and HLA-DR** | 0.8591 (0.7299-0.9883) | 0.0013 |
| **MARS and CD163 and CD192 and HLA-DR** | 0.8545 (0.7222-0.9869) | 0.0015 |
| **MARS and CD200R and CD163 and CD192 and HLA-DR** | 0.8818 (0.7615-1.000) | 0.0006 |

#### Prediction of the presence of carotid plaque

MARS accurately predicted the presence of carotid plaque (C-index 0.94, P=0.0022; Figure 6C) in the 10 participants where it was found, whereas QRISK3 did not significantly discriminate between individuals with and without plaque (Figure 6D). As with cIMT, combining MARS with QRISK3 or other monocyte markers did not improve predictive performance (Table 4).

**Table 4.** C-index for prediction of carotid plaque: MARS alone and in combination with QRISK3 score or with other monocyte biomarkers.

|  | **C-index (95% CI)** | **P value** |
| --- | --- | --- |
| **MARS** | 0.9385 (0.8205-1.000) | 0.0022 |
| **MARS and QRISK3** | 0.9231 (0.8108-1.000) | 0.0031 |
| **MARS and CD200R** | 0.9077 (0.7895-1.000) | 0.0044 |
| **MARS and CD163** | 0.9538 (0.8725-1.000) | 0.0015 |
| **MARS and CD192** | 0.9462 (0.8413-1.000) | 0.0018 |
| **MARS and HLA-DR** | 0.9385 (0.8416-1.000) | 0.0022 |
| **MARS and CD200R and CD163** | 0.9308 (0.8315-1.000) | 0.0026 |
| **MARS and CD200R and CD192** | 0.9385 (0.8326-1.000) | 0.0022 |
| **MARS and CD200R and HLA-DR** | 0.9154 (0.7978-1.000) | 0.0037 |
| **MARS and CD163 and CD192** | 0.9538 (0.8620-1.000) | 0.0015 |
| **MARS and CD163 and HLA-DR** | 0.9462 (0.8604-1.000) | 0.0018 |
| **MARS and CD192 and HLA-DR** | 0.9462 (0.8413-1.000) | 0.0018 |
| **MARS and CD200R and CD163 and CD192** | 0.9538 (0.8315-1.000) | 0.0015 |
| **MARS and CD200R and CD163 and HLA-DR** | 0.9462 (0.8526-1.000) | 0.0018 |
| **MARS and CD163 and CD192 and HLA-DR** | 0.9538 (0.8725-1.000) | 0.0015 |
| **MARS and CD200R and CD163 and CD192 and HLA-DR** | 0.9462 (0.8604-1.000) | 0.0018 |

### 2. VALIDATION STUDY

Baseline demographics of the external validation cohort are presented in Table 5. Given recent evidence questioning the utility of cIMT for risk stratification,^22^ analyses in this cohort focused on carotid plaque rather than cIMT. MARS values were significantly higher in participants with carotid plaque compared with those without plaque (1.19±0.26 vs 0.93±0.22 (P=0.0006). ROC analysis confirmed that MARS reliably discriminated individuals with carotid plaque (C-index 0.81, P<0.0001; Figure 7A). In contrast, China-PAR did not show significant predictive ability (Figure 7B). Although MARS and China-PAR were significantly correlated, this association was weak (r²=0.1031, Figure 7C), again suggesting that MARS reflects biological features not captured by conventional risk calculators.

**Figure 7.**
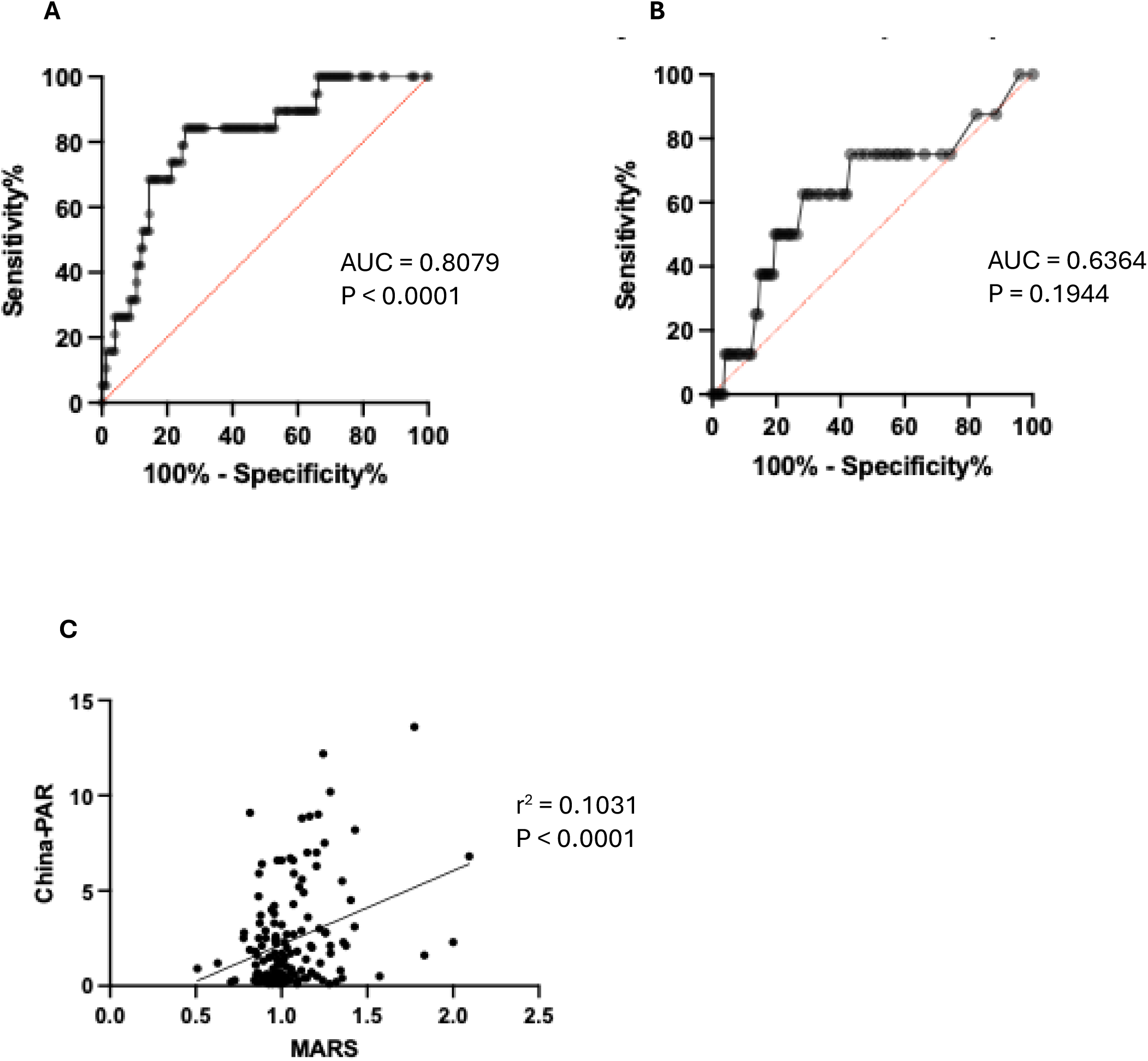
Predictive ability of MARS and China-PAR score for carotid plaque disease in the validation cohort. ROC curves are shown for ability of (**A**) MARS and (**B**) China-PAR to predict the presence of plaque. Also shown is the correlation analysis between MARS and China-PAR **(C)**.

**Table 5.** Demographics of subjects recruited from the Physical Examination Centre at Nanjing Drum Tower Hospital.

|  |  |
| --- | --- |
| **Number (male / female)** | 76/75 |
| **Age (years)** | 47.0±10.5 |
| **Smokers** | 35 |
| **Systolic blood pressure (mmHg)** | 123.5±15.4 |
| **Diastolic blood pressure (mmHg)** | 78.5±10.3 |
| **Creatinine (μmol/L)** | 67.9±15.7 |
| **HbA<sub>1c</sub> (%)** | 5.56±0.75 |
| **Total cholesterol (mmol/L)** | 4.92±0.92 |
| **LDL-cholesterol (mmol/L)** | 3.10±0.81 |
| **HDL-cholesterol (mmol/L)** | 1.41±0.38 |
| **Triglycerides (mmol/L)** | 1.60±0.64 |
| **Haemoglobin (g/L)** | 141.2±17.0 |
| **Total white cell count (× 10<sup>9</sup>/L)** | 5.96±1.51 |
| **Neutrophil count (× 10<sup>9</sup>/L)** | 3.21±1.22 |
| **Monocyte count (× 10<sup>9</sup>/L)** | 0.34±0.12 |
| **Lymphocyte count (× 10<sup>9</sup>/L)** | 1.94±0.52 |
| **Platelet count (× 10<sup>9</sup>/L)** | 238.2±64.2 |

## DISCUSSION

Accurate identification of individuals with subclinical atherosclerosis in order to instigate prophylactic measures including lifestyle modification, lipid lowering and antihypertensive therapies, remains a major unmet need in cardiovascular prevention.^23^ In this study, we show that circulating monocyte phenotype and MPA provide biologically informative signals that strongly associate with carotid atherosclerotic burden in asymptomatic individuals. By integrating these features into a composite index derived from discovery data, we demonstrate improved discrimination of silent carotid disease compared with established cardiovascular risk calculators, with consistent findings across two independent cohorts of individuals diverse in their ethnicity.

Traditional risk calculators, including QRISK3 and China-PAR, estimate cardiovascular risk based on demographic and clinical variables that reflect long-term exposure to established risk factors. While they are effective at a population level, these models do not incorporate biological measures of active vascular inflammation and therefore perform less well when applied to individual patients.^6,7^ They are also limited by other important determinants that may be absent due to limited data or incomplete characterisation. There is, therefore, a clear need for biomarkers that directly identify silent atherosclerotic disease independent of traditional risk factor profiles, not only to improve individual risk stratification but also to inform therapeutic decision-making and trial design. For example, although anti-platelet therapy is well established for secondary prevention, evidence supporting its use in primary prevention remains limited.^24–26^ This likely reflects the design of prior trials, which enrolled broadly defined patient populations without stratification by underlying atherosclerotic burden. Consequently, many participants were unlikely to have biologically relevant disease at baseline, diluting any potential treatment effect. A biomarker capable of accurately identifying silent atherosclerosis in asymptomatic individuals could enable more appropriate selection of trial populations, providing biological enrichment and allowing a clearer assessment of therapeutic benefit in future primary prevention studies. Our data suggest that the monocyte-derived measures examined here may reflect ongoing cellular processes central to atherogenesis, providing complementary information that is not captured by conventional risk models.

The results from our discovery study show that circulating monocyte phenotype as well as MPA levels correlate more closely with cIMT, a well validated surrogate marker of atherosclerotic burden, than QRISK3 score. Our composite index (MARS score) correlates best with cIMT and, furthermore, is a considerably better predictor both of high risk cIMT and of the presence of carotid plaque than is QRISK3. The MARS score integrates opposing monocyte signals, combining increased inflammatory activation with depletion of classical monocytes, thereby capturing a shift in circulating monocyte biology associated with atherosclerosis. This integrated approach outperforms individual monocyte markers and the additional phenotypes examined, suggesting that combining functionally related cellular features yields a more robust biomarker than single parameters alone. The limited improvement observed when adding conventional risk scores or additional monocyte markers further supports the notion that this composite index captures the dominant biological signal relevant to early disease. Validation of MARS in an independent cohort with distinct ethnic and clinical characteristics strengthens the generalisability of our findings. In this population, MARS reliably discriminated individuals with carotid plaque, whereas the regionally appropriate risk calculator, China-PAR, did not. The weak correlation between the two measures suggests that they capture largely independent information, reinforcing the potential value of biologically anchored biomarkers to complement existing risk assessment strategies. Monocytes play a pivotal role in plaque formation, with CD16-expressing subsets exhibiting enhanced interactions with activated endothelium; endothelial adhesion and contact-dependent signalling followed by transmigration into the arterial wall are key early steps in lesion initiation and progression.^27^ MPA provide a circulating readout of platelet–monocyte interactions that amplify this process, where platelet binding to circulating monocytes enhances pro-inflammatory signalling, promotes endothelial adhesion and recruitment, and couples thrombotic activation with vascular inflammation during early atherogenesis.^28^ The associations that we have observed between monocyte subsets, monocyte–platelet aggregates and carotid atherosclerosis are biologically consistent with established experimental and clinical observations in symptomatic disease.^9–14^. Our data now extend these observations to asymptomatic populations and demonstrate that these circulating signatures track with subclinical disease burden.

Imaging modalities such as Duplex ultrasonography and computed tomography (CT) angiography remain the reference standard for identifying atherosclerotic disease. However, their application to large asymptomatic populations is impractical because of limited access to appropriately trained personnel, cost and, in the case of CT, exposure to ionising radiation. A blood-based biomarker that can be applied at scale to identify individuals at high risk of underlying atherosclerosis would therefore allow more targeted and efficient use of vascular imaging. Our findings suggest that MARS provides a sensitive and specific measure of atherosclerotic disease burden that is well suited to this role. Importantly, the degree of individual risk discrimination achieved using this approach exceeds that provided by traditional cardiovascular risk calculators.

We have considered the limitations of our findings. Our discovery cohort was modest in size and the strong associations observed should therefore be interpreted cautiously. However, concerns regarding overfitting are mitigated by the consistency we found across multiple related cellular measures, robustness to combination with other markers and replication in an external validation cohort. We acknowledge that this cross-sectional study focuses on imaging surrogates rather than clinical events, and future prospective studies will be required to determine whether this biomarker predicts cardiovascular events and improves clinical decision-making. Finally, although the use of flow cytometry may be perceived as a limitation for large-scale clinical application, it is now widely embedded in routine diagnostics, and the limited marker panel along with the straightforward staining and gating strategy used in this study are readily compatible with existing clinical platforms and automated acquisition and analysis workflows.

In conclusion, this study demonstrates that circulating monocyte phenotype and MPA analysis provide mechanistically grounded insight into subclinical atherosclerosis that is not captured by conventional risk calculators. A composite monocyte-derived index, MARS, generated from these features identifies silent carotid disease with greater accuracy than established models in two independent asymptomatic populations. These findings support further prospective evaluation of monocyte-based biomarkers as tools to refine individual cardiovascular risk assessment and guide targeted imaging and preventive strategies.

## Data Availability

All data produced in the present study are available upon reasonable request to the authors.

## ACKNOWLEDGEMENTS

We thank Alexander Kerr for his help with running the flow cytometry assays used in the discovery phase of this study.

## AUTHORS’ CONTRIBUTIONS

FA, JG, LNK, SP and SQ performed, and BX, AP and AF conceived and designed, the experimental and clinical work. FA, SQ and AF undertook all statistical analyses. All the authors contributed to writing the manuscript.

